# Blood biochemical parameters as predictors of disease severity and mortality in COVID-19 patients- an updated systematic review and meta-analysis

**DOI:** 10.1101/2021.09.16.21263675

**Authors:** Afsha Majid, Pinki Mishra, Rizwana Parveen, Ram Bajpai, Mohd. Ashif Khan, Nidhi Bharal Agarwal

## Abstract

**Background:** The outbreak of coronavirus disease 2019 (COVID-19) has been rapidly spreading across the globe and poses a great risk to human health. Patients with abnormalities in laboratory parameters are more susceptible to COVID-19. Therefore, we explored the association of blood biochemical parameters with severity and mortality of COVID-19 amongst 3695 patients across seventeen studies.

**Methods:** We searched PubMed, Cochrane library and LitCOVID database until February 28, 2021. Seventeen studies were included in the meta-analysis with 3695 COVID-19 patients.

**Results:** The pooled analysis showed that compared to non-severe group, severe group was characterised by significantly elevated alanine aminotransferase (ALT) (standardised mean difference [SMD]: 0.65, 95% confidence interval [CI]: 0.23 to 1.06; p<0.001, erythrocyte sedimentation rate (ESR) (SMD: 0.55, 95%CI: 0.02 to 1.07 p=0.004) and lymphopenia (SMD: -1.22, 95% CI: -2.15 to -0.30; p<0.01), decreased serum albumin (SMD: -1.60, 95% CI: -2.96 to -0.22 ; p<0.001), creatinine (SMD: 0.54, 95% CI: 0.17 to 0.90; p<0.001), lactate dehydrogenase (LDH)(SMD: -1.54, 95% CI: - 2.27 to -0.80; p=0.002) and haemoglobin (SMD:-0.89, 95% CI: ; p<0.001). Additionally, in the non-survivor group, elevated lactate dehydrogenase (LDH) (SMD: 1.54 95% CI: -2.27 to 0.80; p=0.002), decreased serum albumin (SMD: 1.08, 95% CI: 0.75 to 1.42; p<0.001) were reported. There was no comorbidity which was found to be significant in the severe group.

**Conclusion:** Serum albumin, ALT, ESR, lymphopenia, haemoglobin, and leucocytosis can reflect the severity of COVID-19, while the LDH, leucocytosis and albumin can be considered as risk factor to higher mortality.

**Summary:** Our manuscript discusses the various blood biochemical markers as potential predictors of disease severity and mortality in COVID-19 patients. The timely detection of these parameters can help in providing appropriate course of treatment and reducing the mortality rate in the patients. We have found an association between the blood biochemical markers and disease severity and mortality in COVID-19 patients. Serum albumin, alanine aminotransferase (ALT), Erythrocyte Sedimentation Rate (ESR), lymphopenia, hemoglobin, and leukocytosis can reflect the severity of the disease, while the LDH, leukocytosis and albumin can be considered as risk factor to higher mortality.

## 1. Introduction

Coronavirus disease 2019 (COVID-19) is a global pandemic caused by the severe acute respiratory syndrome coronavirus 2 (SARS-CoV-2). The first case was reported in Wuhan (China), in December 2019. The number of cases and the associated mortality has increased dramatically (1). SARS-CoV-2 causes respiratory infections and is like Severe Acute Respiratory Syndrome Virus (SARS-CoV) (genome sequence-80 –90 per cent similar) and Middle East Respiratory Syndrome Virus (MERS-CoV) in terms of clinical symptoms. As per WHO (World Health Organisation) Coronavirus Update Report of February 14, 2021, there have been 108, 153, 741 confirmed cases of COVID-19 and 2, 381, 295 deaths worldwide (2).

The clinical spectrum of COVID-19 ranges from mild to severe form of the disease (3). Mild symptoms include fever, cough followed by sputum production, and fatigue; while severe symptoms include acute respiratory distress (ARDS) syndrome, and acute cardiac injury along with multiple organ failure (4).

There are various laboratory parameters such as lymphocytes, leukocytes, haemoglobin, liver enzymes, albumin, inflammatory markers and procalcitonin that are found to be deranged during the SARS-CoV-2 infection (5,6). As per previous literature, liver enzymes such as ALT, AST and bilirubin are found to be elevated in severe form of the illness due to possible liver dysfunction by the virus (1,4,7). Albumin levels have been found to decrease in severe illness (8–10). The inflammatory markers such as C-reactive protein were also found to be elevated in cases of severe infection (11). Procalcitonin, which is found to be elevated in bacterial infections (12) was found to be in the normal range in SARS-CoV-2 infection. As the production of procalcitonin increases during bacterial infection (13), there is simultaneous increased levels of IL-1β, tumour necrosis factor (TNF) and IL-6, but in case of viral infections these are inhibited by interferon-γ release. Thus, it can be postulated that procalcitonin will only increase if there is a bacterial co-infection in the patient along with COVID -19 otherwise it will be within the range (12). Creatinine is primarily excreted by the kidneys and abnormally high levels of creatinine is an indication of renal insufficiency which is a common complication in COVID-19 patients (14).

Several comorbidities such as hypertension, diabetes, chronic heart disease (CHD), chronic kidney disease (CKD) and chronic obstructive pulmonary disorder (COPD) are known to be prevalent in the severe group (15–17). However, more evidence needs to be gathered about their individual prevalence. There are various complications such as ARDS, pneumonia, kidney injury, septic shock as well as secondary infections which are also known to be related to the severe form of the illness and also related to the mortality of the COVID-19 patients (18,19). As the cases of COVID-19 infection are rapidly increasing, conduct of a comprehensive and detailed research providing better insights in this domain has become essential. Not only it will help in the early diagnosis of the severe form of the disease but also it will help in formulating better treatment modalities and risk stratification in COVID-19 patients.

Although, similar systematic reviews have been published previously (20,21). However, our systematic review has several merits including: a) most updated database search b) broader research question and, c) included additional parameters and outcomes in our meta-analysis which gives a detailed insight into the association of these parameters with disease severity and mortality in COVID-19 patients.

Therefore, in the present meta-analysis, we aimed to understand the association between laboratory parameters and disease severity and mortality of COVID-19 patients. This will not only help broaden the horizon but also lead to the optimisation of the use of resources for the population at risk.

## 2. Materials and Methods

“Preferred Reporting Items for Systematic Review and Meta-Analysis (PRISMA) guidelines for systematic reviews (22) and meta-analysis of observational studies in epidemiology (MOOSE) guidelines (23) were followed for designing, conduct and reporting this systematic literature review.” The protocol of this review has been registered with PROSPERO (registration number CRD42020206741).

### 2.1 Data sources and searches

We searched PubMed/Medline, Cochrane Central Register of Controlled Trials and Clinical Trial Registry-India until February 28, 2021 using the keywords “laboratory” OR “clinical”, OR “lab parameters”, “comorbidities”, “clinical outcome” AND “coronavirus 2019” OR “COVID-19” OR “2019nCoV-2”, OR “SARS CoV-2”. We also searched grey literature using Google Scholar and reference list of eligible articles with the aim of identifying additional potential eligible studies.

### 2.2 Inclusion and exclusion criteria

Eligible studies were cross-sectional, case-control, cohort and case series reporting defined groups and extractable data on laboratory findings in confirmed COVID-19 patients were included. The editorials, reviews, letters, meta-analysis, consensus and case reports, studies not reported in English language were excluded from the study. First author (AM) searched data and screened article for eligibility. Senior author (PM) double checked all the included articles and any dispute was resolved by consensus.

### 2.3 Quality assessment

Two reviewers (AM and PM) assessed the quality of data in the included studies using the National Institute of Health (NIH) quality assessment tools (24). We preferred the NIH tool because it is comprehensive and widely accepted for an exhaustive assessment of data quality. We rated the overall quality of included studies as good, fair and poor, and incorporated them in the meta-analysis results.

### 2.4 Data extraction

Data were inputted into a standardized data extraction table (Excel) and independently checked by a second reviewer (PM) for accuracy. The following variables were extracted: name of the first author, year of publication, study design, gender, age, number of patients in severe and non-severe groups, days of hospitalization along with comorbidities of all the patients and clinical outcomes in terms of death and discharge. If not mentioned, the mean and standard deviation were extrapolated by median, sample size and interquartile range (IQR) (25). The severity of disease was defined according to Diagnosis and Treatment Plan of COVID-19 issued by National Health Commission, China (7^th^ edition) as mild, common, severe, and critical based on the clinical symptoms (26).

### 2.5 Data synthesis

We performed an exploratory meta-analysis to understand the magnitude and direction of effect estimate. Continuous outcomes are presented using standardised mean difference (SMD) due to substantial variability in study designs and 95% confidence intervals (CIs). We interpreted the effect size using Cohen rule of thumb with SMD greater than or equal to 0.2 representing a small effect, SMD greater than or equal to 0.5 a moderate effect, and SMD greater than or equal to 0.8 a large

Effect (27) For dichotomous outcomes, risk ratios (RRs) were calculated and presented with respective 95% CIs. Mantel-Haenszel random-effects meta-analysis using DerSimonian and Laird method was used to pool ORs (28). Heterogeneity between studies was assessed using the χ^2^-based Cochran’s Q statistic (p<0.1 considered as the presence of heterogeneity) and I-squared (*I*^*2*^) statistics (>50% representing moderate heterogeneity) (28). If any cell value was zero, then we added 0.5 to each cell to calculate risk ratio (29). Publication bias was not assessed as a total number of studies were less than ten in primary lab outcomes (28).

## 3. Results

### Search results

The systematic search yielded a total of 792 publications. Out of 792 articles, 225 were found using search terms “laboratory findings and COVID-19”, 108 articles were found using the keywords “laboratory parameters and COVID -19”, 120 studies were found using the keywords “comorbidities and COVID-19”. After removing duplicates, out of 453 studies, 110 articles were excluded because they were review articles (n=30), did not report data on COVID-19 disease (n=36), did not provide laboratory data on COVID-19 patients with or without severe or without the proper categorization of the patients (n=34), or were editorial material (n=6) and 4 articles were excluded as they were in Chinese full text and could not be translated into English. Five additional studies were identified from the reference list of selected articles. All studies reported laboratory values measured at admission or earliest time point on hospitalization. Except for one study in which the classification of disease severity was unclear and hence it was not included in the meta-analysis. Thus, the meta-analysis included 17 studies meeting the inclusion criteria (Figure 1).

**Fig.1.**
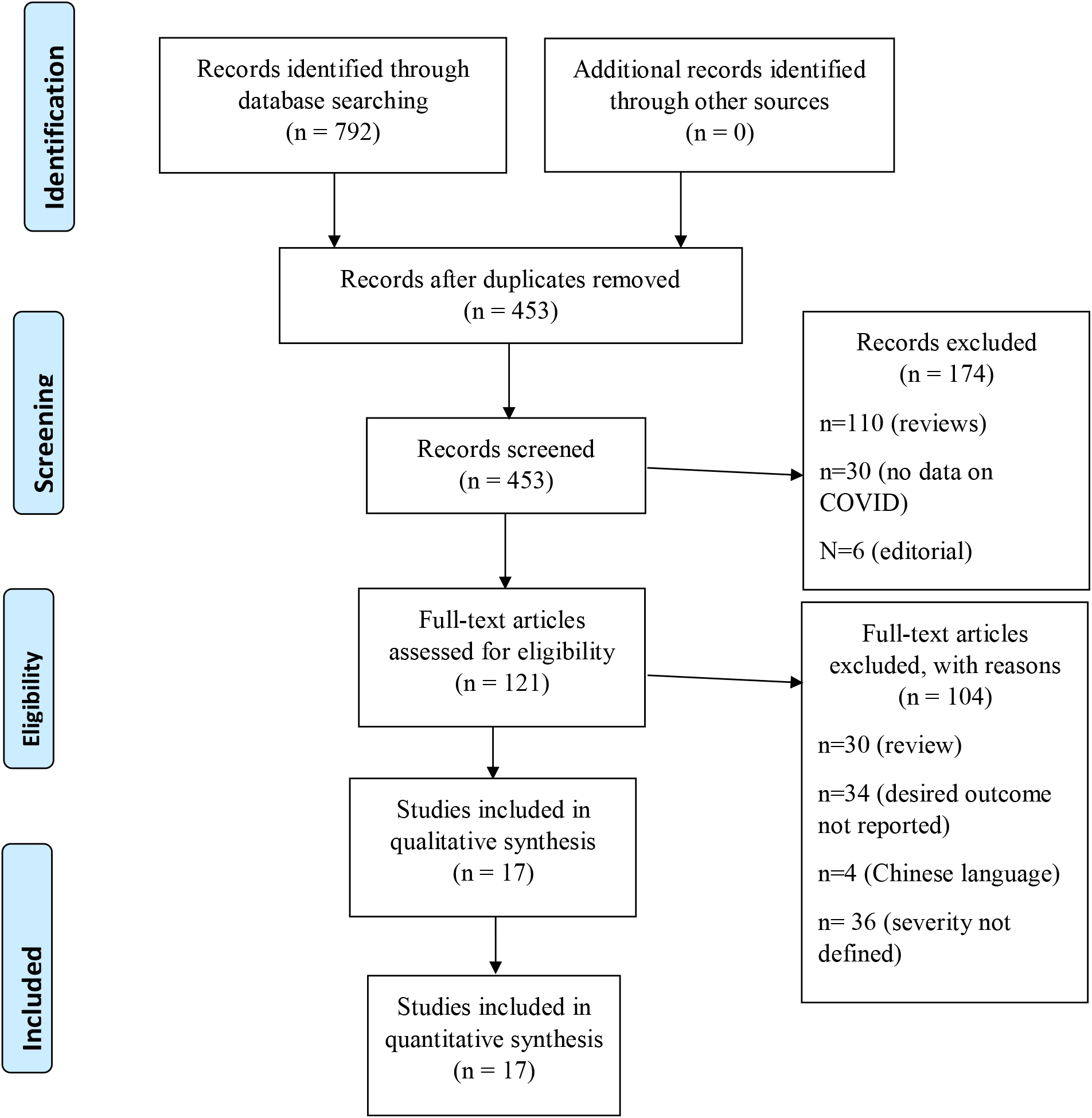
Flow diagram of the number of studies screened and included in the meta-analysis.

### 3.1 Study Characteristics

All the included studies were found to be conducted in China. Out of 17 included studies, 11 studies reported data in severe and non-severe groups, 6 studies reported data in terms of patient’s survival with survivor and non-survivor groups. Among the seventeen included studies, five were cross-sectional studies (4,30–33), four were cohort studies (34) while two were case-series. The included studies enrolled a total of 3695 patients, including 1884 males and 1811 females. The baseline characteristics of the subjects included in these studies are provided in Table-1.

**Table 1:**
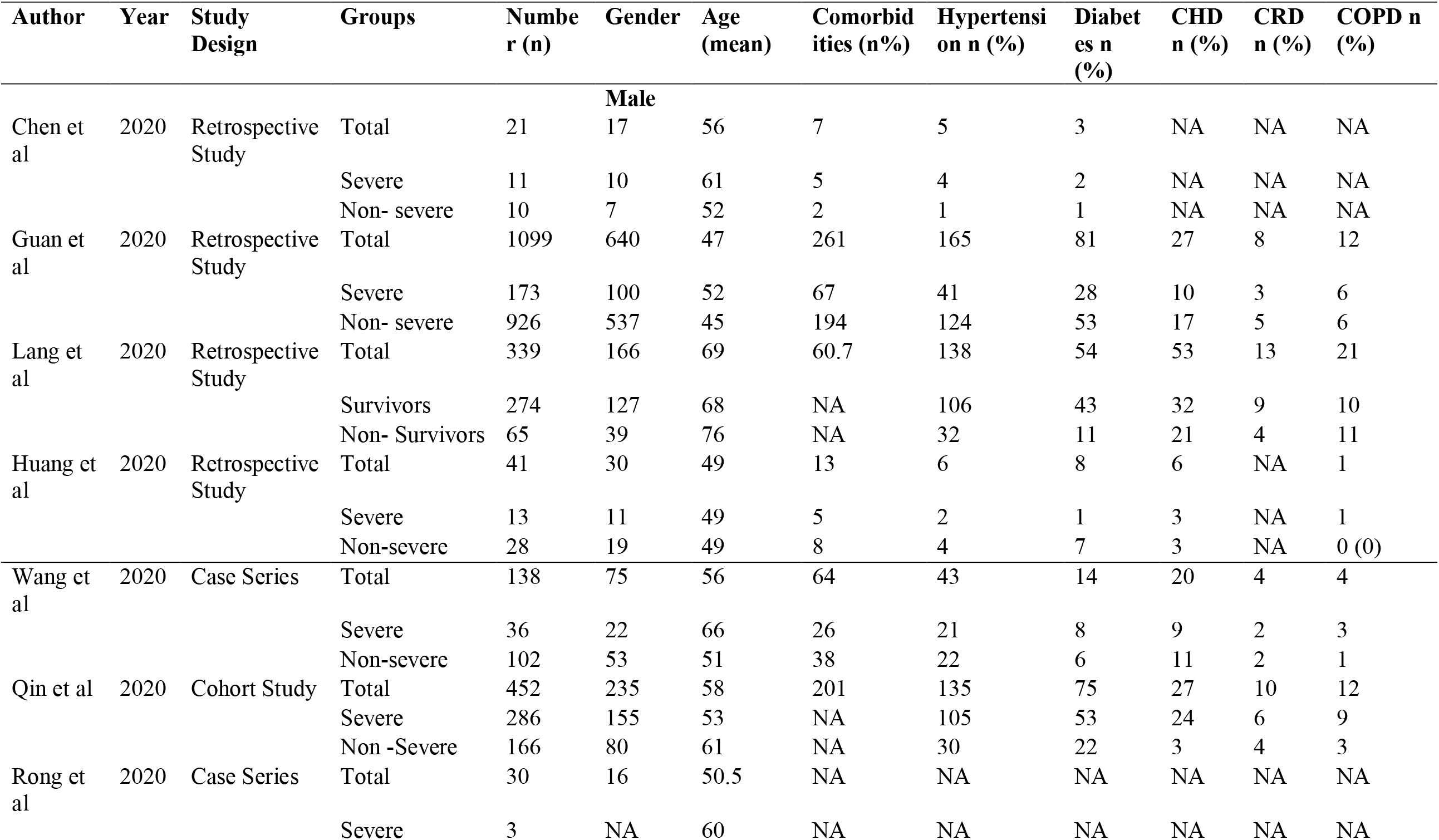
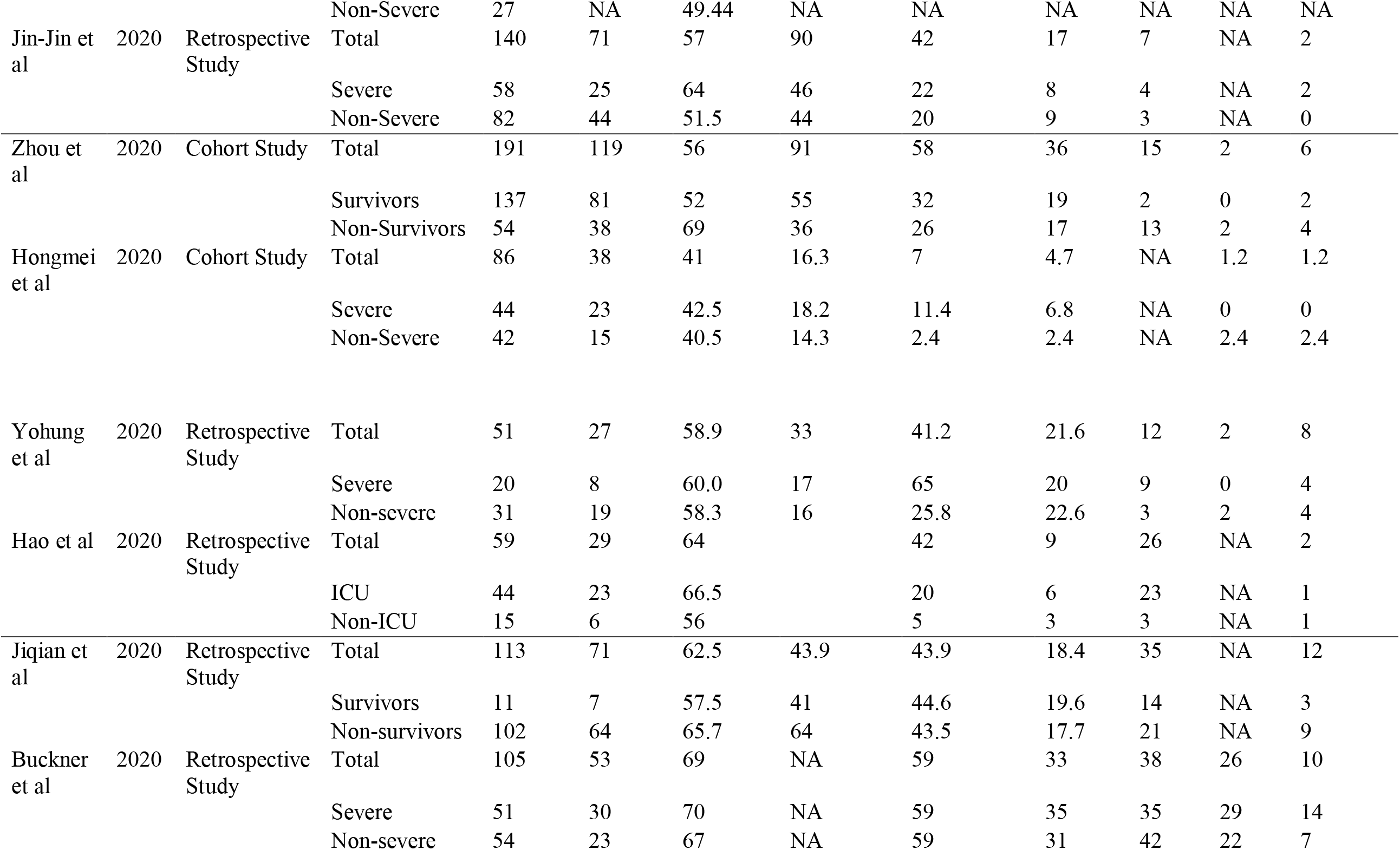
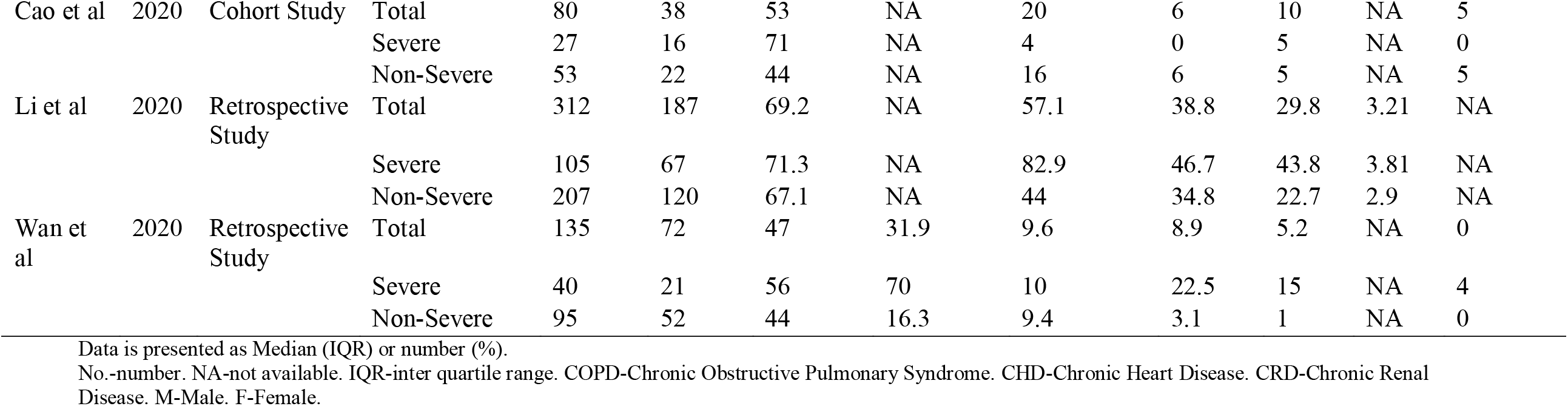
Baseline characteristics of the subjects

The laboratory parameters of patients were taken into consideration along with the major comorbidities such as hypertension, diabetes, CHD, CRD. Complications in the patient’s post-treatment such as shock, acute respiratory distress syndrome (ARDS), kidney disease as well as secondary infections were also assessed.

### 3.2 Quality Assessment

We assessed the quality of data included in the studies using the NIH quality assessment tools. The quality assessment indicated that most included studies were of acceptable quality. All the studies clearly stated the research question or objective, the study population was clearly defined, and all subjects were selected from the same or similar population. The detailed result of the quality assessment is provided in *Supplementary File*.

### 3.3 Laboratory Findings

Regarding the laboratory findings, the parameters that were significantly elevated in the severe group were ALT (p<0.001) and ESR (p<0.001), while the albumin (p<0.001), haemoglobin (p<0.001) and leukocyte count (leucopoenia) (p<0.001) were found significantly decreased in the severe group as compared to the non-severe group. However, no significant difference was found in other parameters such as creatinine, procalcitonin, AST, C-reactive protein, lymphocyte count and LDH. Levels of LDH, albumin and leucocyte count were found associated with the survival of the patient (Table 2).

**Table 2:**
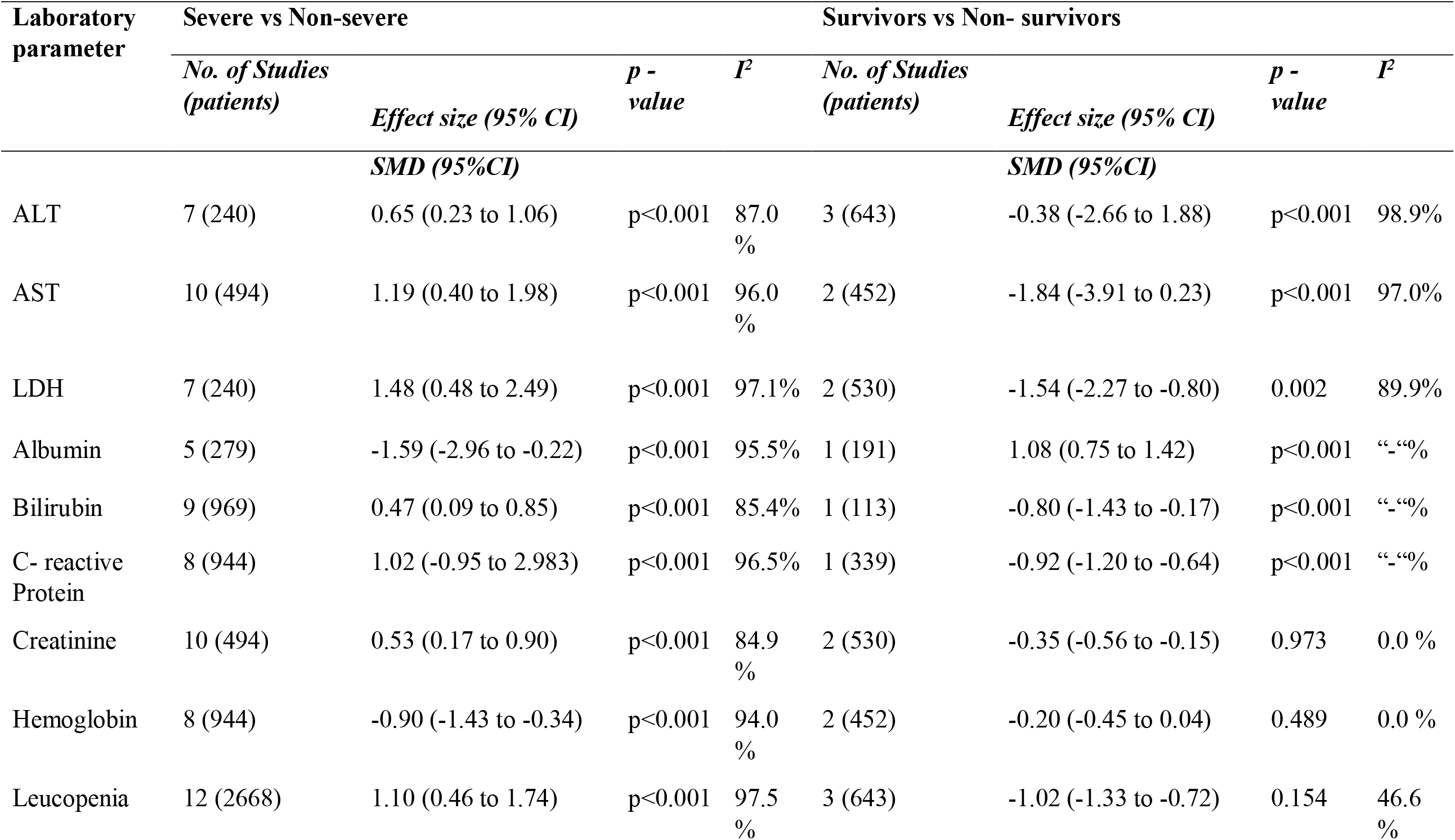
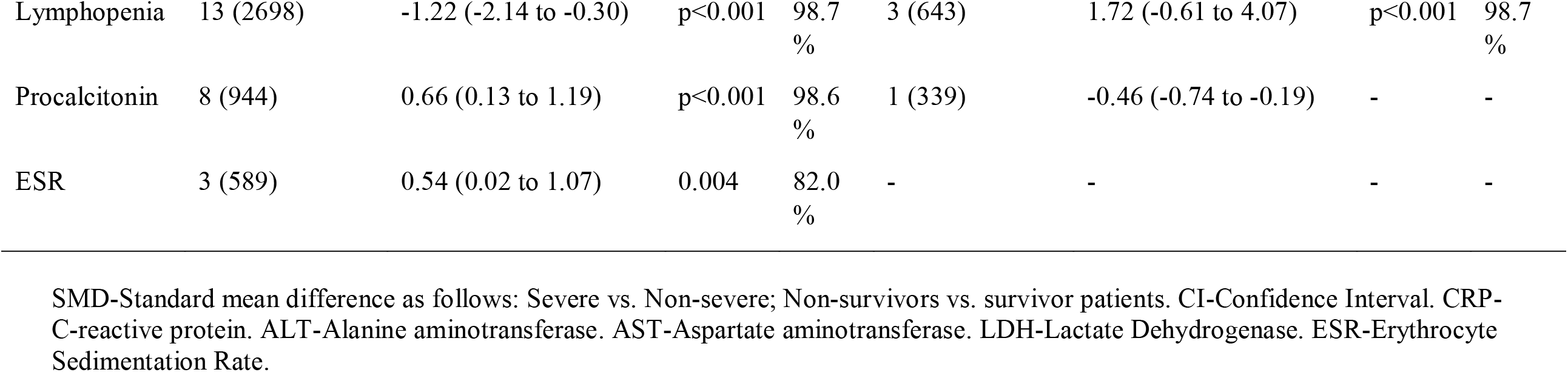
Results of meta-analysis comparing laboratory abnormalities in COVID-19 patients with and without severe illness and mortality.

### 3.4 Comorbidities

The prevalence of Hypertension (p=0.005) and Chronic Heart Disease (CHD) (p=0.001) and Chronic Obstructive Pulmonary Syndrome (COPD) (p=0.001) was found significantly higher in the severe group while there was no significant difference in the prevalence of Diabetes, Chronic Renal Disease (CRD) in the two groups. None of the above-mentioned comorbidities was related to the mortality of the patient (Table 3).

**Table 3:**
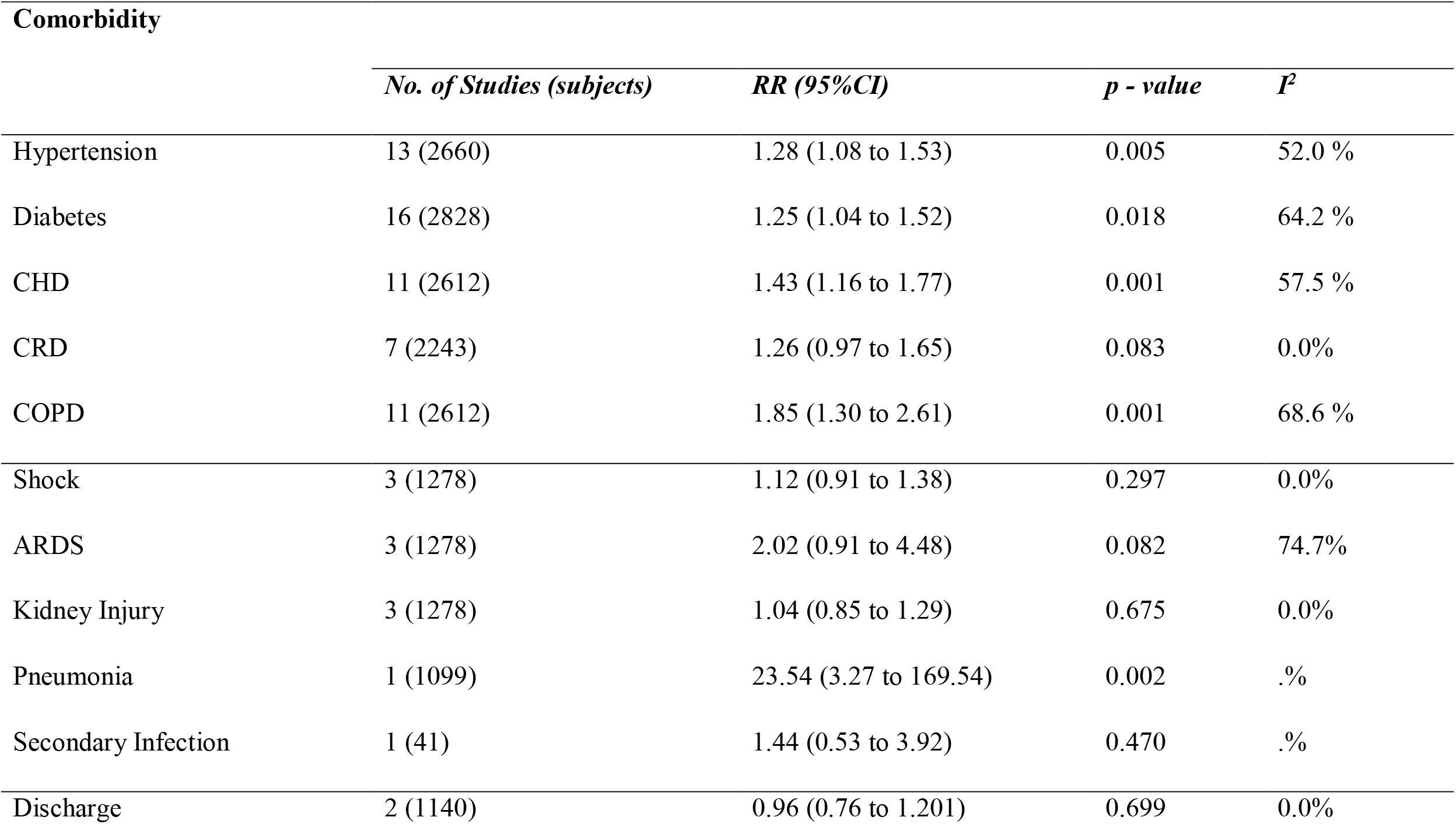
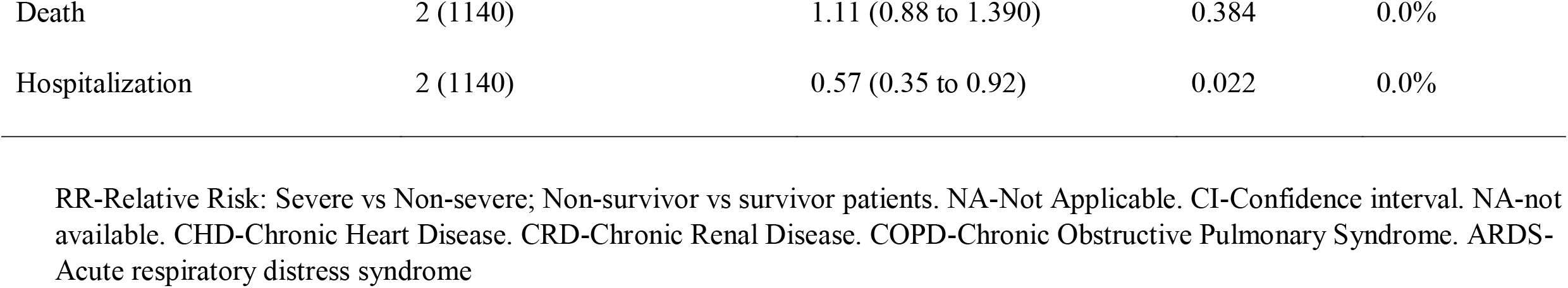
Results of meta-analysis comparing co-morbidities, complications, and clinical outcome in COVID-19 patients with and without severe illness and mortality.

| **Comorbidity** |  |  |  |  |
| --- | --- | --- | --- | --- |
|  | *No. of Studies (subjects)* | *RR (95%CI)* | *p - value* | *I<sup>2</sup>* |
| Hypertension | 13 (2660) | 1.28 (1.08 to 1.53) | 0.005 | 52.0 % |
| Diabetes | 16 (2828) | 1.25 (1.04 to 1.52) | 0.018 | 64.2 % |
| CHD | 11 (2612) | 1.43 (1.16 to 1.77) | 0.001 | 57.5 % |
| CRD | 7 (2243) | 1.26 (0.97 to 1.65) | 0.083 | 0.0% |
| COPD | 11 (2612) | 1.85 (1.30 to 2.61) | 0.001 | 68.6 % |
| Shock | 3 (1278) | 1.12 (0.91 to 1.38) | 0.297 | 0.0% |
| ARDS | 3 (1278) | 2.02 (0.91 to 4.48) | 0.082 | 74.7% |
| Kidney Injury | 3 (1278) | 1.04 (0.85 to 1.29) | 0.675 | 0.0% |
| Pneumonia | 1 (1099) | 23.54 (3.27 to 169.54) | 0.002 | .% |
| Secondary Infection | 1 (41) | 1.44 (0.53 to 3.92) | 0.470 | .% |
| Discharge | 2 (1140) | 0.96 (0.76 to 1.201) | 0.699 | 0.0% |
| Death | 2 (1140) | 1.11 (0.88 to 1.390) | 0.384 | 0.0% |
| Hospitalization | 2 (1140) | 0.57 (0.35 to 0.92) | 0.022 | 0.0% |

RR-Relative Risk: Severe vs Non-severe; Non-survivor vs survivor patients. NA-Not Applicable. CI-Confidence interval. NA-not available. CHD-Chronic Heart Disease. CRD-Chronic Renal Disease. COPD-Chronic Obstructive Pulmonary Syndrome. ARDS-Acute respiratory distress syndrome

### 3.5 Complications

Out of the several complications studied, pneumonia (p=0.02) was the only significant complication in the severe group while there was no significant difference found in complications such as shock, ARDS, kidney injury and secondary infection between the two groups. None of the above-mentioned complications was associated with mortality of the patient (Table 3).

### 3.6 Clinical Outcome

The rate of hospitalisation (p=0.022) was more in the severe group while there was no significant difference in the death and discharge rates among the two groups (Table 3).

## 4. Discussion

Over the last one year, more than 111,762,965 cases of COVID-19 have been confirmed in China and other countries in Asia, Pacific, Europe, Africa, and the America. Clinical and laboratory, and factors associated with evolution of the disease and outcomes, constitute critical knowledge that should be cautiously studied when a new infectious disease arises. COVID-19 has a wide spectrum of disease severity ranging from asymptomatic to symptomatic, mild to severe and critical nature of the illness (35). In this meta-analysis, we aimed to assess the association between the laboratory parameters along with comorbidities and complications with disease severity and mortality in 3695 COVID-19 patients. (36). The laboratory parameters that were observed in this meta-analysis were the elevated ALT and ESR while decreased levels of albumin and haemoglobin were noted. However, the levels of other parameters such as bilirubin, LDH, CRP, AST, lymphocyte count, leucocyte count and procalcitonin were not found significantly different in the two groups. Data from the 2002-2003 outbreak indicate that SARS may be associated with lymphopenia, leukopenia, and, elevated levels of Lactate Dehydrogenase (LDH), Alanine transaminase (ALT), Amino Aspartate Transaminase (AST), and creatine kinase (37,38) but they are not significantly seen, nor consistently reported, in COVID-19 studies and cases. The mortality was found to be associated with the LDH, albumin, leukopenia and leucocytosis across two studies (32,39).

As per previous studies, hypertension and diabetes have been reported to be the most prevalent comorbidities found in COVID-19 patients. In a study in China, out of 13 patients, hypertension was found in 5 (27.8%) and diabetes was found in 3 (16.7%) (16,40,41). In contrary to this, hypertension was the most prevalent comorbidity observed in the severe group while there was no significant difference in the prevalence of diabetes, CHD, CRD, and COPD between the two groups. None of the above-mentioned comorbidity had an association with the mortality of the patient.

Complications that were commonly observed in the severe group was pneumonia while the occurrence of shock, kidney injury, ARDS and secondary infections were similar in the two groups and only ARDS and kidney injury were related to the mortality of the patients. The rate of hospitalisation was more in the severe group while there was no significant difference found in the rates of death and discharge between the two groups. Our results showed a deviation from the usual findings due to heterogeneity between the individual studies and small number of studies included in the meta-analysis. Thus, a more comprehensive analysis with a larger sample size needs to be done so that we can have a clear picture of the correlation of laboratory abnormalities and clinical parameters with the severity of the disease. More such studies are required to elucidate the risk factors for disease severity and death.

There are several limitations that needs to be mentioned. There was heterogeneity amongst individual studies because of which there was a deviation of some of our results from usual findings. Additionally, case-series were included in the present meta-analysis. Although we did an extensive search, we may have inadvertently missed relevant studies. Exclusion of studies in languages other than English (ie. Chinese) may have resulted in missing of relevant studies. Certain parameters such as IL-6 ad IL-10 which are strong indicators of cytokine storm were not included in this meta-analysis due to lack of available data as not all parameters were reported in each patient. As most of the articles were published in Chinese, findings should interpret with caution, thereby ensuring the generalizability of the results. Also, not all studies included all the desired parameters.

## 5. Conclusion

COVID-19 has a wide spectrum of severity. The detection of these laboratory as well as clinical parameters can assist in the timely diagnosis of such patients. They can be associated with disease severity and mortality along with deciding the course of action for COVID-19 positive patients and reducing the mortality rates in the patients. The patients of liver dysfunction with abnormal levels of liver enzymes and comorbidities such as hypertension are more prone to severe form of COVID-19 infection and therefore, special attention should be given to such patients. However, more such studies with a greater sample size needs to be conducted to get a better insight into the role of liver enzymes in the prognosis of COVID-19.

## Data Availability

The author confirms that the data supporting the findings of the study are available within the article [and/ or its supplementary material].

## 5. Acknowledgement

We are thankful to all the authors who contributed significantly towards the completion of this study.

## Preferred Reporting Items for Systematic reviews and Meta-Analyses extension for Scoping Reviews (PRISMA-ScR) Checklist

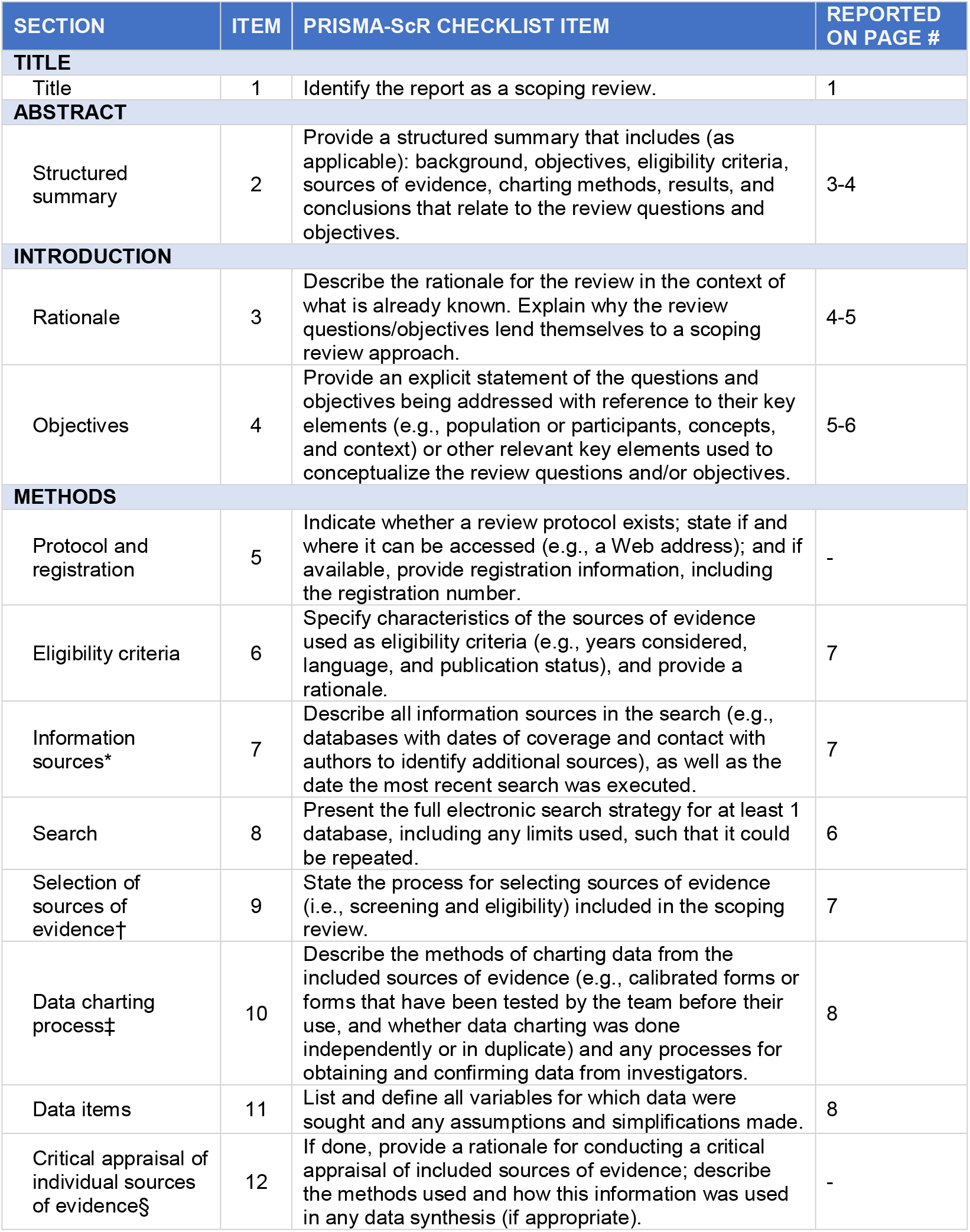
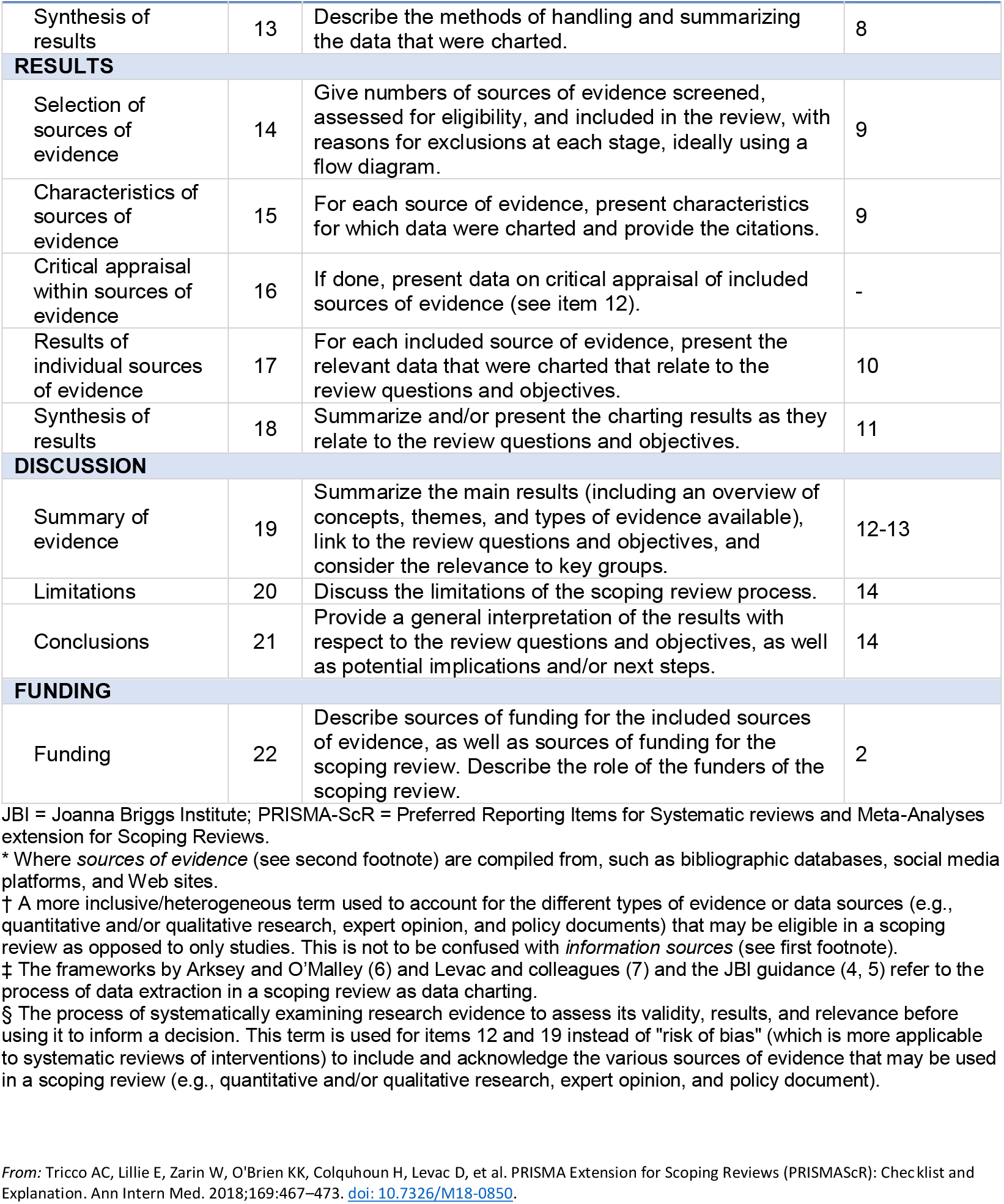

## Notes

### Competing Interest Statement

The authors have declared no competing interest.

### Funding Statement

This research did not receive any specific grant from funding agencies in the public, commercial, or not-for-profit sectors.

