## Supplementary material for "Blood biochemical parameters as predictors of disease severity and mortality in COVID-19 patients- an updated systematic review and meta-analysis"

Supplementary file

1. **Quality assessment of Included Studies Using NIH tool of quality assessment**

*Quality assessment of cohort and cross-sectional studies*

| **Criteria** | **Chen et al 2020** | **Guan et al 2020** | **Lang et al 2020** | **Huang et al 2020** | **Qin et al 2020** | **Jin-Jin et al 2020** | **Zhou et al 2020** | **Hongmei et al** | **Yohung et al** | **Hao et al** | **Jiqian et al** | **Buckner et al** | **Cao et al** | **Li et al** | **Wan et al** |
| --- | --- | --- | --- | --- | --- | --- | --- | --- | --- | --- | --- | --- | --- | --- | --- |
| Was the research question or objective in this paper clearly stated? | Yes | Yes | Yes | Yes | Yes | Yes | Yes | Yes | Yes | Yes | Yes | Yes | Yes | Yes | Yes |
| Was the study population clearly specified and defined? | Yes | Yes | Yes | Yes | Yes | Yes | Yes | Yes | Yes | Yes | Yes | Yes | Yes | Yes | Yes |
| Was the participation rate of eligible persons at least 50%? | Yes | Yes | Yes | Yes | Yes | Yes | Yes | Yes | Yes | Yes | No | No | Yes | No | Yes |
| Were all the subjects selected or recruited from the same or similar populations (including the same time period)? Were inclusion and exclusion criteria for being in the study prespecified and applied uniformly to all participants? | Yes | Yes | Yes | Yes | Yes | Yes | Yes | Yes | Yes | Yes | Yes | Yes | Yes | Yes | Yes |
| Was a sample size justification, power description, or variance and effect estimates provided? | NA | NA | NA | NA | NA | NA | NA | NA | NA | NA | NA | NA | NA | NA | NA |
| For the analyses in this paper, were the exposure(s) of interest measured prior to the outcome(s) being measured? | Yes | Yes | Yes | Yes | Yes | Yes | Yes | Yes | Yes | Yes | Yes | Yes | Yes | No | Yes |
| Was the timeframe sufficient so that one could reasonably expect to see an association between exposure and outcome if it existed? | Yes | Yes | Yes | Yes | Yes | Yes | Yes | Yes | Yes | Yes | Yes | Yes | Yes | Yes | Yes |
| For exposures that can vary in amount or level, did the study examine different levels of the exposure as related to the outcome (e.g., categories of exposure, or exposure measured as continuous variable)? | NA | NA | NA | NA | NA | NA | NA | NA | NA | NA | NA | NA | NA | NA | NA |
| Were the exposure measures (independent variables) clearly defined, valid, reliable, and implemented consistently across all study participants? | Yes | Yes | Yes | Yes | Yes | Yes | Yes | Yes | Yes | Yes | Yes | Yes | Yes | Yes | Yes |
| Was the exposure(s) assessed more than once over time? | NA | NA | NA | NA | NA | NA | NA | NA | NA | NA | NA | NA | NA | NA | NA |
| Were the outcome measures (dependent variables) clearly defined, valid, reliable, and implemented consistently across all study participants? | Yes | Yes | Yes | Yes | Yes | Yes | Yes | Yes | Yes | Yes | Yes | Yes | Yes | Yes | Yes |
| Were the outcome assessors blinded to the exposure status of participants? | NA | NA | NA | NA | NA | NA | NA | NA | NA | NA | NA | NA | NA | NA | NA |
| Was loss to follow-up after baseline 20% or less? | NA | NA | NA | NA | NA | NA | NA | NA | NA | NA | NA | NA | NA | NA | NA |
| Were key potential confounding variables measured and adjusted statistically for their impact on the relationship between exposure(s) and outcome(s)? | Yes | Yes | Yes | Yes | Yes | Yes | Yes | Yes | Yes | Yes | Yes | Yes | Yes | Yes | Yes |
| Quality Rating (CD, cannot determine; NA, not applicable; NR, not reported | Good | Good | Good | Good | Good | Good | Good | Good | Fair | Fair | Fair | Good | Good | Good | Fair |

| **Criteria** | **Wang et al 2020** | **Rong et al 2020** |
| --- | --- | --- |
| Was the study question or objective clearly stated? | Yes | Yes |
| Was the study population clearly and fully described, including a case definition? | Yes | Yes |
| Were the cases consecutive? |  |  |
| Were the subjects comparable? | Yes | Yes |
| Was the intervention clearly described? | NA | NA |
| Were the outcome measures clearly defined, valid, reliable, and implemented consistently across all study participants? | Yes | Yes |
| Was the length of follow-up adequate? | Yes | Yes |
| Were the statistical methods well-described? | Yes | Yes |
| Were the results well-described? | Yes | Yes |
| Quality Rating (CD, cannot determine; NA, not applicable; NR, not reported | Good | Good |

*Quality assessment of case-series*
